## Supplementary material for "Estimating the effectiveness of non-pharmaceutical interventions against COVID-19 transmission in the Netherlands"

* corresponding author

### S1 Estimation of relative fitness of COVID-19 variants

For SARS-CoV-2 the basic reproduction number increased over time due to the emergence of new, more transmissible variants. Here, we assume that the growth advantages of all variants before the Omicron variant were completely attributable to a higher transmissibility (that is, to a higher $R_0(t)$). To estimate the transmissibility by variant, we use the genomic surveillance data that consists of numbers of randomly sampled isolates by variant per week in the Netherlands. A multinomial logistic regression model is fitted to the data to estimate the relative growth rates $\lambda_v$ for each variant $v$ relative to wildtype (i.e., $\lambda_{\mathrm{wildtype}} = 0$) [1]. The transmissibility $F_v$ of variant $v$ is found by solving the Euler-Lotka equation [2]:

$$F_v \int_0^\infty \exp(-\lambda_v \tau) g_v(\tau) d\tau = 1,$$

with $g_v(t)$ the generation interval distribution of variant $v$. Assuming a gamma distributed generation interval with mean $\mu_v$ and shape $\alpha_v$, yields [3]:

$$F_v = \left(1 + \frac{\lambda_v \mu_v}{\alpha_v}\right)^{\alpha_v}.$$

We take all pre-Omicron variants to have a mean generation interval of 4 days and a shape parameter of 4 [4]. Finally, the overall transmissibility over time $F(t)$ is the average of the variant transmissibilities weighted to their fitted proportions $p_v(t)$ (Fig. 1):

$$F(t) = \sum_v F_v \; p_v(t).$$

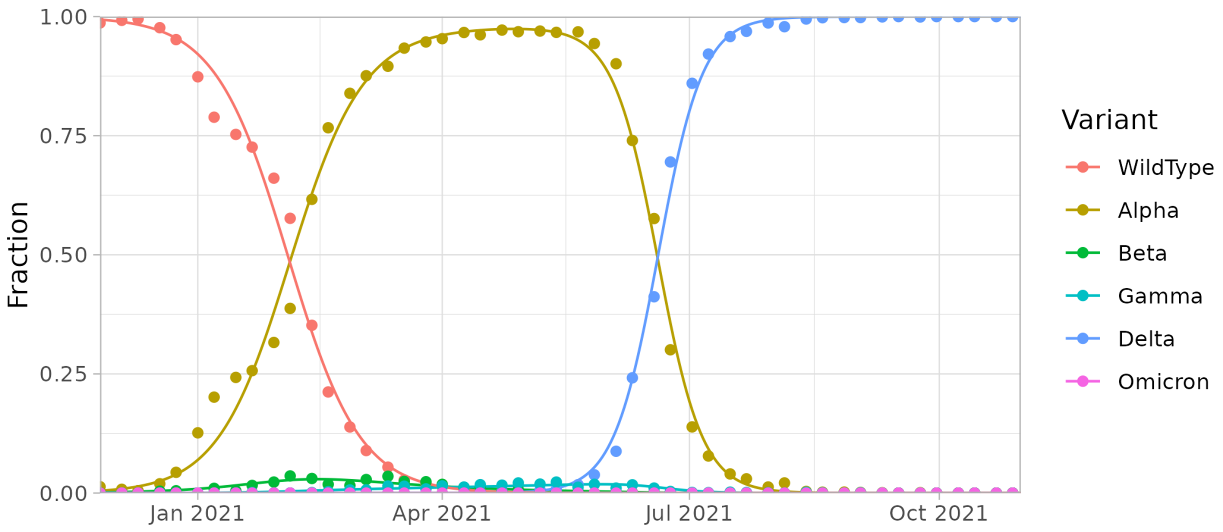


Figure 1: Fraction of SARS-CoV-2 variants over time from genomic surveillance data, fitted (lines) and data (points).

### S2 Reconstruction of population immunity

#### S2.1 Definition of immunity classes

To reconstruct the population immunity we subdivide each age group in classes depending on the infection, vaccination and immune status (Fig. 2), where we assume that vaccination is independent of the infection status. In each age group $j$ a fraction $q_j(t)$ is infected at least once, which we infer from serological surveys, but only a fraction $\phi_{\text{inf},j}(t)$ is still naturally immune. Also, a fraction $v_j(t)$ is vaccinated, which we know from vaccination data of the primary series, but only a fraction $\phi_{\text{vac},j}(t)$ is still immune due to vaccination.

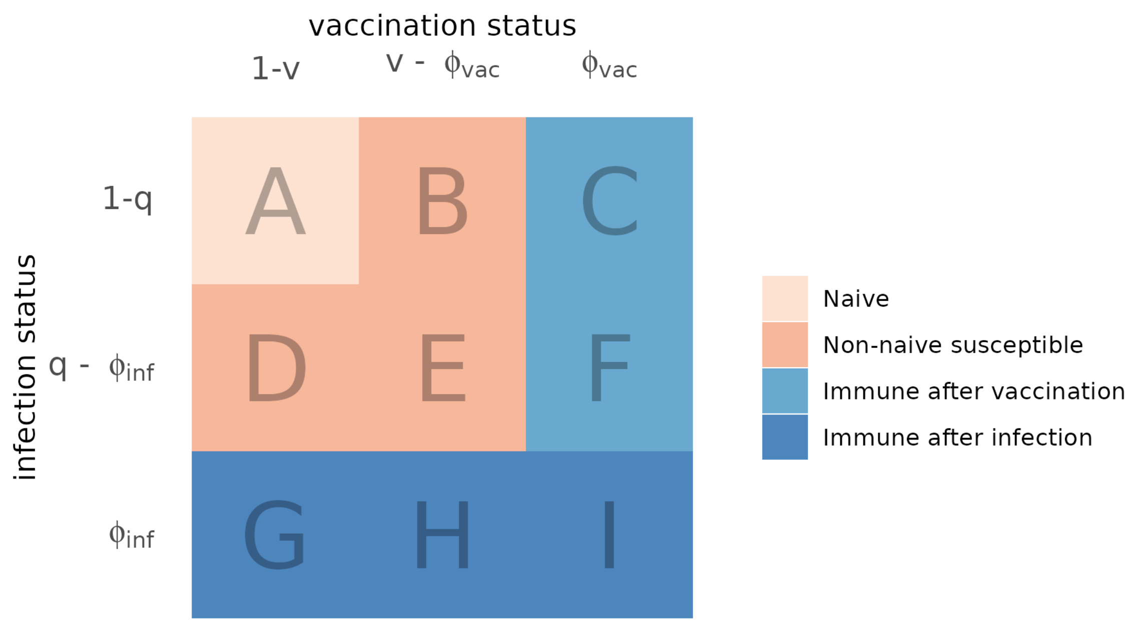


Figure 2: Population immunity classes for each age group of which fraction $q$ is infected at least once, $v$ vaccinated, $\phi_{\text{inf}}$ immune after infection and $\phi_{\text{vac}}$ immune after vaccination. The letters denote the different combinations of infection and vaccination status, of which I has in fact immunity after infection and vaccination.

The immune fraction $\phi_j(t)$ in age group $j$ (blue classes in Fig. 2) is calculated by:

$$\phi_j(t) = \phi_{\text{inf},j}(t) + \phi_{\text{vac},j}(t) - \phi_{\text{inf},j}(t)\phi_{\text{vac},j}(t).$$

For the immune fraction in the full population $\phi(t)$ that is used to calculate the effectiveness, we take the population average of the immune fractions per age group:

$$\phi(t) = \frac{\sum_j n_j(t)\phi_j(t)}{\sum_j n_j(t)}.$$

For simplicity, we assume all susceptibles within the same age group are equally susceptible, and when infected they are equally infectious. In this way, the fraction of new infections that are reinfections, $p_{\text{reinf},j}(t)$, is

$$p_{\text{reinf},j}(t) = \frac{q_j(t) - \phi_{\text{inf},j}(t)}{1 - \phi_{\text{inf},j}(t)},$$

and the fraction of new infections that are breakthrough infections, $p_{\text{break},j}(t)$, is

$$p_{\text{break},j}(t) = \frac{v_j(t) - \phi_{\text{vac},j}(t)}{1 - \phi_{\text{inf},j}(t)}.$$

Below, we explain how we imputed the cumulative infected fractions in each age group, which assumptions we made on immunity waning, and how we calculated the immune fractions.

#### S2.2 Cumulative infected fraction

The nationwide serological Pienter-Corona survey was held every 2-6 months from April 2020 onwards [5]. For each participant the round in which they were first infected was determined. Only participants from the first two rounds (April 2020 and June 2020) were included, as the status of later enrolled participants was unknown. Participants were weighted by sex, age, ethnic background and degree of urbanisation to match the distribution of the general Dutch population, and the fraction of participants that was infected at least once was determined per survey round and per age group using the R-package *survey* yielding the cumulative infected fraction $Q_{j,r}$ in age group $j$ and survey round $r$ with a 95% confidence interval. This fraction reflects all naive infections that occurred up to 11 days before the round, which is the assumed period between infection and a detectable serological response.

To fill in the blanks between survey rounds, we use reported case data. For each reported case the age group and a date of statistics is known. This date of statistics is either the symptom onset or the date of the test-positive result. Assuming 2 days between symptom onset and test-positive result, we construct the notified case data $C_j(t)$ denoting the number of cases in age group $j$ who had their (imputed) symptom onset on day $t$. We shift these time series with an incubation period of 5 days, and we take the weekly moving average to straighten out day-of-week effects, yielding the incidence $c_j(t)$ of notified cases as a fraction of age group size $n_j(t)$ on the day of infection $t$:

$$c_j(t) = \frac{1}{7\, n_j(t)} \sum_{k=-3}^{k=3} C_j\,(t + 5 + k).$$

The increase of the cumulative infected fraction from one survey round to the next, was caused by the cumulative true incidence between those two survey rounds shifted by 11 days. When we assume that the reporting rate in an age group is constant between two rounds, the true incidence is proportional to the observed incidence $c_j(t)$. We use this to calculate a scaling factor $m_j(t)$ for age group $j$ at day $t$:

$$m_j(t) = \sum_r \frac{\left(Q_{j,r} - Q_{j,r-1}\right)\left[u\left(t - \theta_{j,r-1} + 11\right) - u\left(t - \theta_{j,r} + 11\right)\right]}{\sum_{\tau=\theta_{j,r-1}-10}^{\theta_{j,r}-11} c_j\,(\tau)},$$

with $\theta_{j,r}$ is the median date of survey round $r$ in age group $j$ and $u(x)$ defined as a step function which is 0 if $x < 0$ and 1 if $x \geq 0$. The resulting scaling factors are piecewise constant. In the final analysis the survey rounds 1 and 3 were omitted; the first round because the serological test was still being developed and the third round because it was too close to the second round. The cumulative infected fraction $q_j(t)$ is the cumulative scaled incidence:

$$q_j(t) = \sum_{\tau=0}^{t} c_j(\tau)\, m_j(\tau).$$

#### S2.3 Immunity loss due to waning

We assume the following functional forms for the immunity waning profiles after infection $\pi_{\mathrm{inf}}(t)$ and vaccination $\pi_{\mathrm{vac}}(t)$:

$$\begin{aligned}
\pi_{\mathrm{inf}}(t) &= 0 && t < 0\\
&= 1 && 0 \le t < 42\\
&= \exp\big(-\omega_{\mathrm{inf}}(t-42)\big) && t \ge 42
\end{aligned}$$

and

$$\begin{aligned}
\pi_{\mathrm{vac}}(t) &= 0 && t < 0\\
&= \mathrm{VE}_{\max}\left(\frac{t+14}{28}\right) && 0 \le t < 14\\
&= \mathrm{VE}_{\max} && 0 \le t < 42\\
&= \mathrm{VE}_{\max}\exp\big(-\omega_{\mathrm{vac}}(t-42)\big) && t \ge 42,
\end{aligned}$$

where $\mathrm{VE}_{\max}$ is the maximal vaccine effectiveness against infection, and $\omega_{\mathrm{inf}}$ and $\omega_{\mathrm{vac}}$ are the exponential waning rates after infection and (the second dose of) vaccination, the values of which depend on the fast, medium or slow waning scenario. These waning profiles are identical for all age groups and variants.

#### S2.4 Calculation of immune fraction after vaccination

The fraction $\phi_{\mathrm{vac},j}(t)$ of age group $j$ that is immune due to vaccination is found by convolution of the vaccination incidence with the vaccine immunity waning profile:

$$\phi_{\mathrm{vac},j}(t) = \sum_{\tau<t} \big(v_j(t) - v_j(t-1)\big)\, \pi_{\mathrm{vac}}(t-\tau),$$

where $v_j(t)$ the vaccinated fraction in age group $j$ at time $t$.

#### S2.5 Calculation of immune fraction after infection

Calculating the immune fraction after infection can be found by convolution of the infection incidence with the infection immunity waning profile. However, the infection incidence is made up by first infections that are observed and reinfections that are not observed. The fraction of reinfections needs to be inferred. Note that we can disregard the vaccination status in this derivation, as it is independent of the infection status.

We start by determining the infection pressure $\lambda_j(t)$ in age group $j$ at time $t$ in those who have never been infected before, dividing the fraction of new infections by the never infected fraction:

$$\lambda_j(t) = \frac{q_j(t+1) - q_j(t)}{1 - q_j(t)}.$$

This infection pressure is the same for those who were infected before but who lost their immunity, because we assume that all susceptibles are equally susceptible irrespective of their infection history. We first define $\xi_j(\tau, t)$ as the fraction of infected persons in age group $j$ at time $t$ with their most recent infection at time $\tau$. At this time the susceptible fraction that has been infected before at time $\tau$ is

$$\xi_{\text{sus},j}(\tau, t) = \sum_{\tau<t} \xi_j\,(\tau, t)\big(1 - \pi_{\text{inf}}(t - \tau)\big),$$

of which a fraction of $\lambda_j(t)$, the infection pressure, will be reinfected:

$$\xi_{\text{reinf},j}(\tau, t) = \lambda_j(t)\xi_{\text{sus},j}(\tau, t).$$

The infected fraction is updated for the next time step by summing the first infections and reinfections at time $t$:

$$\xi_j(t, t+1) = q_j(t+1) - q_j(t) + \sum_{\tau<t} \xi_{\text{reinf},j}\,(\tau, t)$$

and subtracting the reinfections to account for the fact that their most recent infection time has been changed:

$$\xi_j(\tau, t+1) = \xi_j(\tau, t) - \xi_{\text{reinf},j}(\tau, t)$$

Finally, the immune fraction is found by convolution with the infection immunity waning profile:

$$\phi_{\text{inf},j}(t) = \sum_{\tau<t} \xi_j\,(\tau, t)\pi_{\text{inf}}(t - \tau).$$

### S3 Reconstructed population immunity for fast and slow waning rates

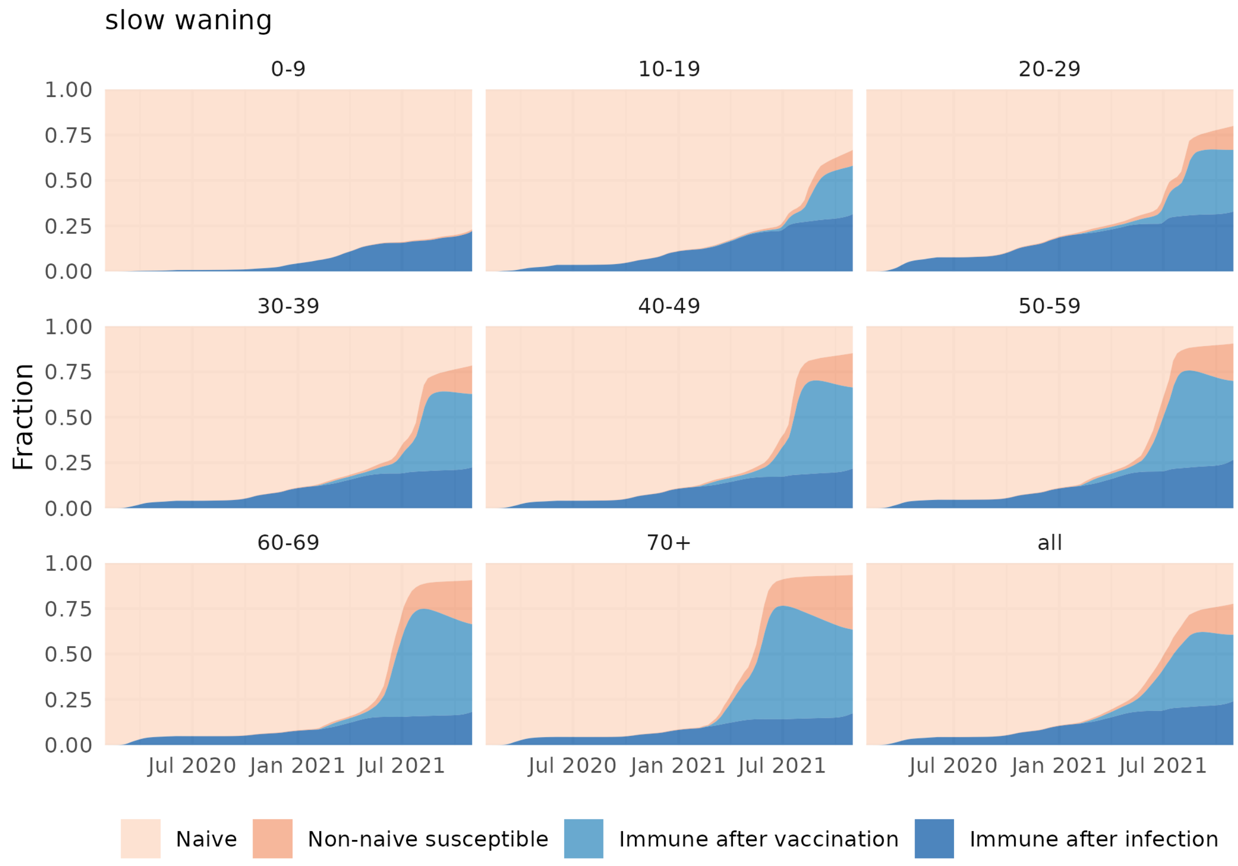


Figure 3: Reconstructed population immunity by age group (and overall) over time (1 February 2020 - 31 October 2021), assuming slow waning rates for natural and vaccine immunity and a high vaccine effectiveness. The population is stratified in a naive class that has never been infected nor vaccinated, a class immune after infection, a class immune after vaccination and a non-naive susceptible class that lost its natural or vaccine immunity.

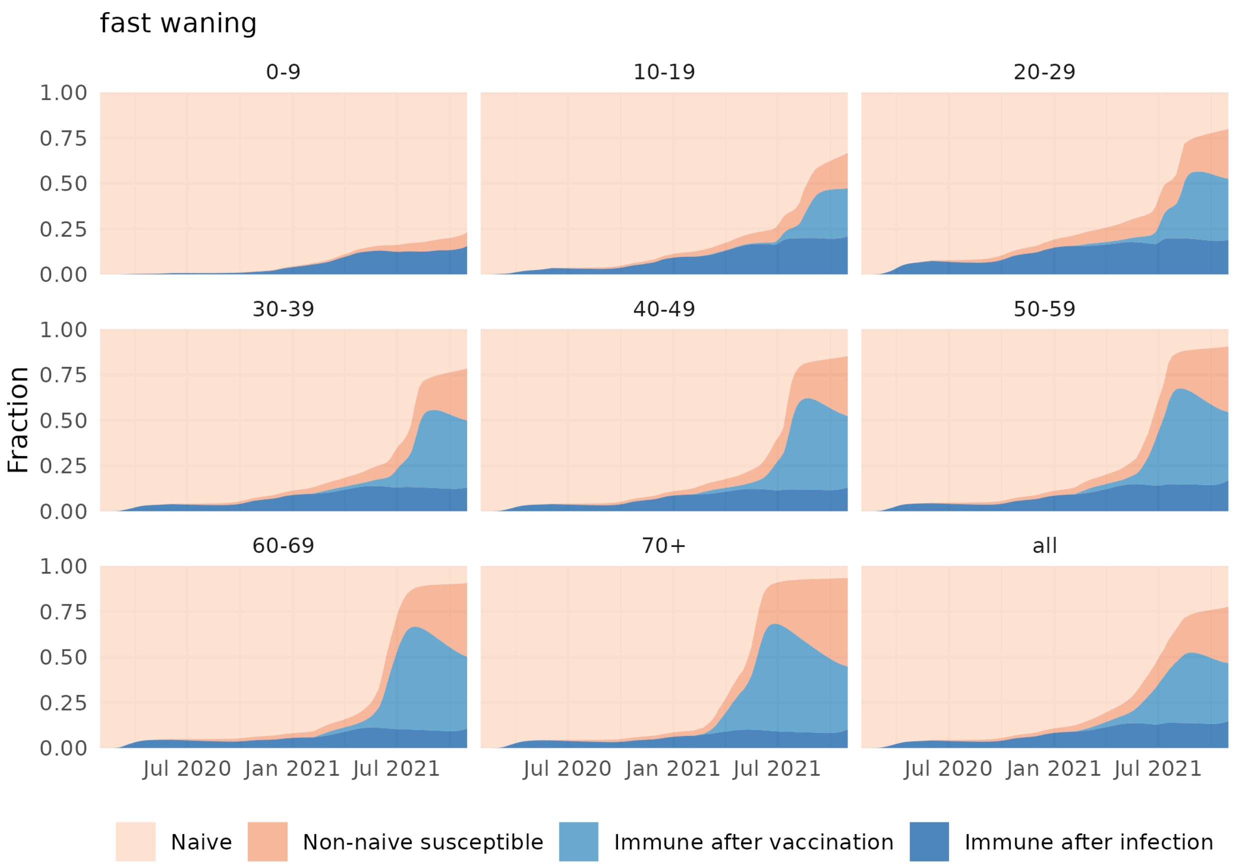


Figure 4: Reconstructed population immunity by age group (and overall) over time (1 February 2020 - 31 October 2021), assuming fast waning rates for natural and vaccine immunity and a low vaccine effectiveness. The population is stratified in a naive class that has never been infected nor vaccinated, a class immune after infection, a class immune after vaccination and a non-naive susceptible class that lost its natural or vaccine immunity.

### S4 Fraction of reinfections and breakthrough infections in new infections

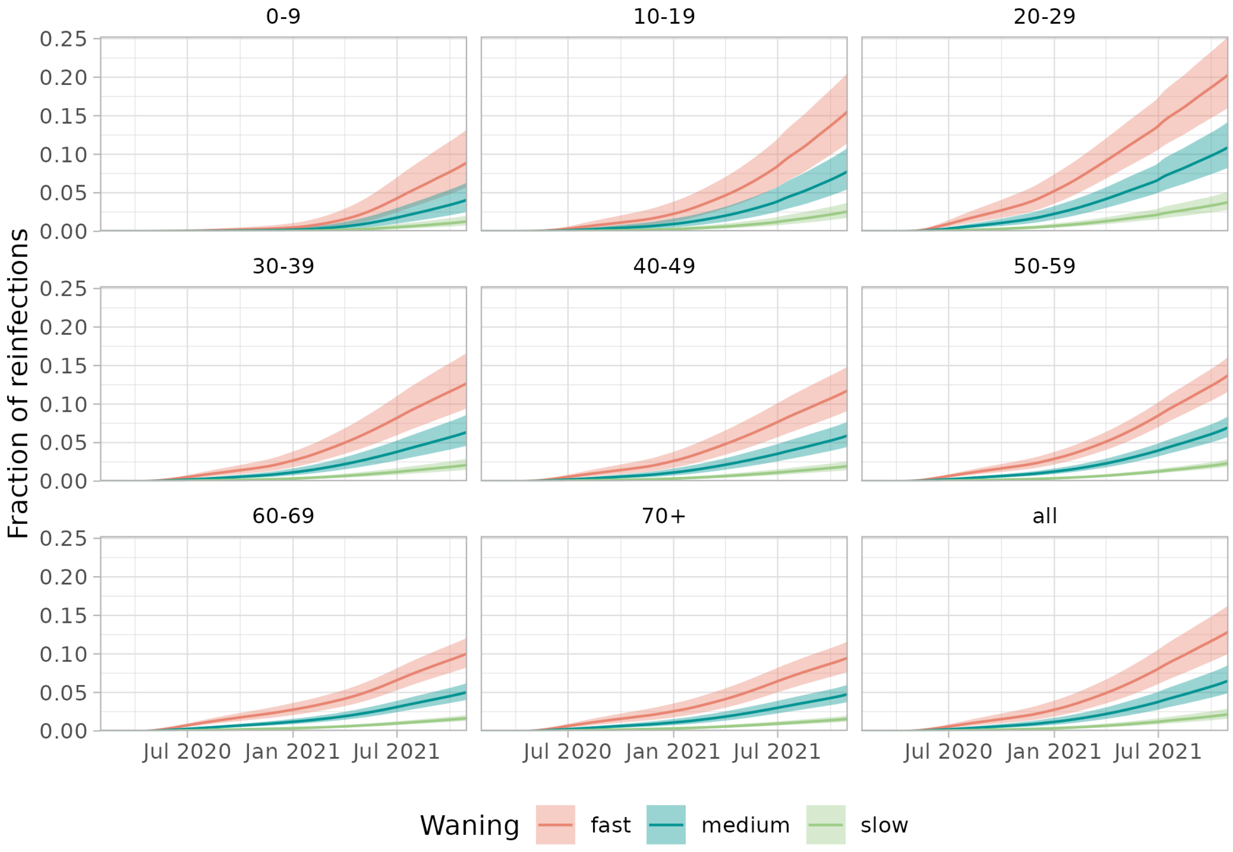


Figure 5: Reconstructed fraction of reinfections in new infections by age group (and overall as presented in main text), assuming slow, medium or fast waning.

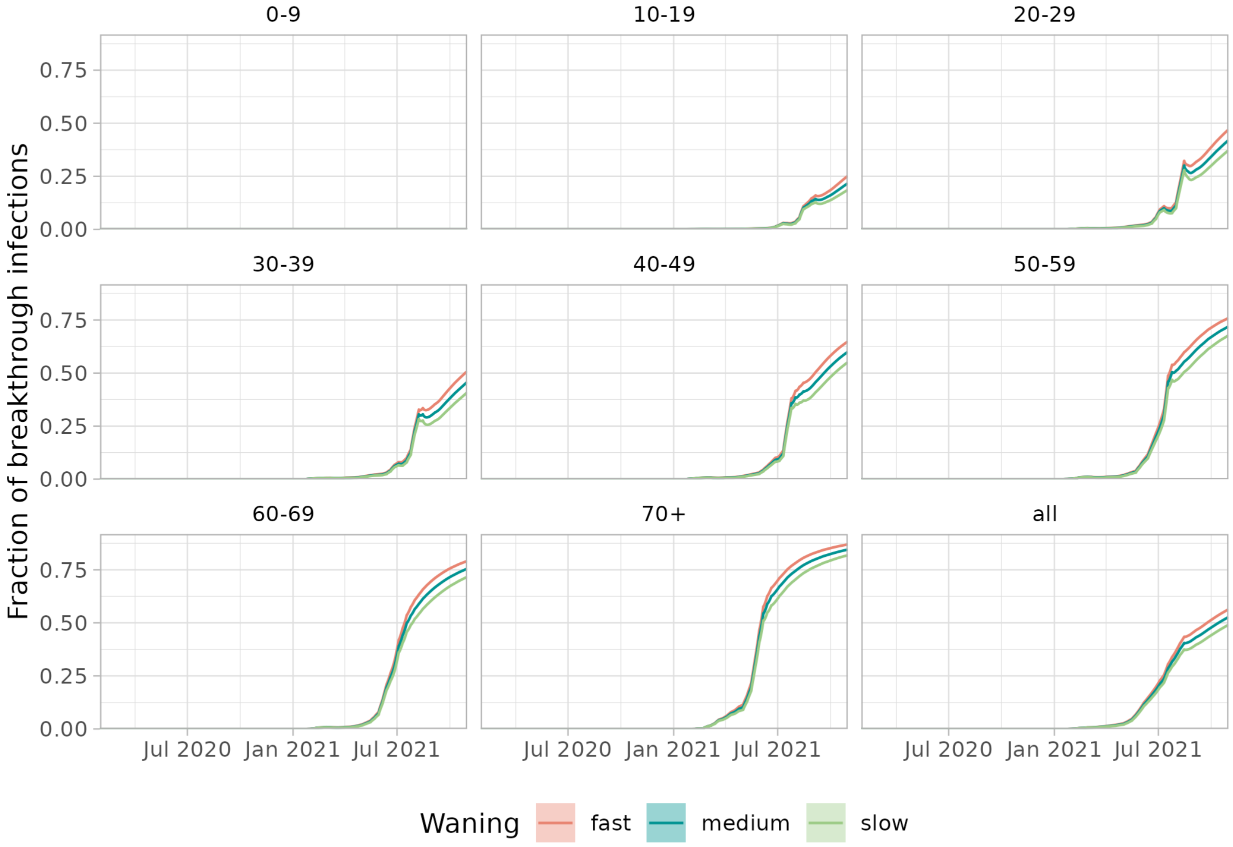


Figure 6: Reconstructed fraction of breakthrough infections in new infections by age group (and overall as presented in main text), assuming slow, medium or fast waning.
